# Effectiveness and cost-effectiveness of community-based TB screening algorithms using Computer-Aided Detection (CAD) technology alone compared to CAD combined with point-of-care C-reactive protein testing: protocol for a paired screen-positive trial

**DOI:** 10.1101/2025.01.16.25320687

**Authors:** Aita Signorell, Alastair van Heerden, Irene Ayakaka, Bart K.M. Jacobs, Marina Antillon, Fabrizio Tediosi, Anna Verjans, Curdin Brugger, Niklaus D Labhardt, Shannon Bosman, Mashaete Kamele, Mamatlakeng Keitseng, Thandanani Madonsela, Johanna Kurscheid, Josephine Muhairwe, Alfred Kipyegon Keter, Keelin Murphy, Bram van Ginneken, Tinne Gils, Bulemba Katende, Rediet Fikru Gebresenbet, Rahel Milena Erhardt, Thomas Zoller, Fiona Vanobberghen, Tracy R. Glass, Lutgarde Lynen, Klaus Reither

## Abstract

**Introduction:** Tuberculosis (TB) remains a significant public health challenge in many African communities, where underreporting and underdiagnosis are prevalent due to barriers in accessing care and inadequate diagnostic tools. This is particularly concerning in hard-to-reach areas with a high burden of TB/HIV co-infection, where missed or delayed diagnoses exacerbate disease transmission, increase mortality, and lead to severe economic and health consequences. To address these challenges, it is crucial to evaluate innovative, cost-effective, community-based screening strategies that can improve early detection and linkage to care.

**Methods and Analysis:** We conduct a prospective, community-based, diagnostic, pragmatic trial in communities of the Butha Buthe District in Lesotho and the Greater Edendale area of Msunduzi Municipality, KwaZulu-Natal in South Africa to compare two strategies for population-based TB screening: Computer-Aided Detection (CAD) technology alone (*CAD4TBv7 approach*) vs. CAD combined with point-of-care C-reactive protein (CRP) testing (*CAD4TBv7-CRP approach*). Following a chest X-ray, CAD produces an abnormality score, which indicates the likelihood of TB. Score thresholds informing the screening logic for both approaches were determined based on the World Health Organization’s (WHO) target product profile for a TB screening test. CAD scores above a threshold pre-specified for the *CAD4TBv7 approach* indicate confirmatory testing for TB (Xpert MTB/RIF Ultra). For the *CAD4TBv7-CRP approach*, a CAD score within a pre-defined window requires the conduct of the second screening test, CRP, while a score above the respective upper threshold is followed by Xpert MTB/RIF Ultra. A CRP result above the selected cut-off also requires a confirmatory TB test. Participants with CAD scores below the (lower) threshold and those with CRP levels below the cut-off are considered screen-negative. The trial aims to compare the yield of detected TB cases and cost-effectiveness between two screening approaches by applying a paired screen-positive design. 20,000 adult participants will be enrolled and will receive a posterior anterior digital chest X-ray which is analysed by CAD software.

**Ethics and dissemination:** The protocol was approved by National Health Research Ethics Committee in Lesotho (NH-REC, ID52-2022), the Human Sciences Research Council Research Ethics Committee (HSRC REC, REC 2/23/09/20) and the Provincial Health Research Committee of the Department of Health of KwaZulu-Natal (KZ_202209_022) in South Africa and from the Swiss Ethics Committee Northwest and Central Switzerland (EKNZ, AO_2022-00044). This manuscript is based on protocol version 4.0, 19 January 2024. Trial findings will be disseminated through peer-reviewed publications, conference presentations and through communication offices of the consortium partners and the project’s website (https://tbtriage.com/).

**Trial registration:** ClinicalTrials.gov (NCT05526885), South African National Clinical Trials Register (SANCTR; DOH-27-092022-8096).

**Article summary:** *Strengths and limitations of this study:*

- This pragmatic trial will constitute the first comparative evaluation of two screening approaches for pulmonary TB using CAD4TB alone or in combination with point-of-care CRP.
- Results from this study will help in identifying the most (cost-)effective strategy for community-based TB screening.
- Pragmatic inclusion criteria and trial delivery will allow an approximation to the real world.

## Introduction

The large pool of undetected tuberculosis (TB) in African communities remains a major public health concern because it results in prolonged infectiousness and disease transmission, higher risk of suffering and death because of delayed onset of treatment, serious health sequelae and catastrophic financial consequences [1,2]. According to the World Health Organization (WHO), about 30% of the incident cases in 2022 were underreported or underdiagnosed, either because of barriers in accessing care or inadequate diagnosis [3]. In particular, underserved people in hard-to-reach communities with high TB/HIV burden are at high risk of missed or delayed diagnosis [4]. Therefore, active systematic screening for TB remains an essential part of TB control in these communities as well as in people with HIV (PWHIV) or diabetes mellitus, or among migrants, miners or prisoners [5].

Community-based TB screening with strong linkage to care has the potential to offer a cost-effective, impactful diagnostic strategy for people living in rural areas of high TB/HIV burden countries, if they are carried out with adequate coverage and intensity [6–8]. Provider-initiated systematic screening is used to distinguish individuals with a high likelihood of having TB from people who are highly unlikely to have TB. Positive screening tests are not meant to be diagnostic and should lead to further evaluation and confirmation [5]. A screening strategy, in which an inexpensive, rapid screening test is performed first, and if positive, followed by a more expensive, time-consuming confirmatory assay, could substantially reduce diagnostic costs, improve early access to diagnosis and improve outcomes [9]. Screening tests should perform as rule-out tests with a high sensitivity, while desired specificity is a trade-off between cost and operational efficiency [10]. As defined at a WHO consensus meeting on target product profiles (TPPs), a TB screening test should be: (a) non-sputum based; (b) easy to use; rapid; and (d) accurate (sensitivity > 90%, specificity > 70%) [5,11].

Computer-aided detection (CAD) of TB-related abnormalities on chest radiography (e.g. CAD4TB, Delft Imaging Systems, NL) and the point-of-care C-reactive protein assay (CRP; e.g. LumiraDx, UK) are two tests which have potential to become widely used screening tests for TB [11–17].

CAD4TB is a digital chest X-ray analysis software using artificial intelligence to generate an abnormality score, which indicates the likelihood of TB. CRP is a cytokine-induced acute phase protein, elevated in infectious diseases such as TB that can be measured with fingerprick blood tests. In 2021, the WHO conditionally recommended that CAD software programmes may be used in place of human readers for the interpretation of digital CXR in screening for TB and that CRP may be used to screen for TB disease among adults and adolescents living with HIV [5].

We are conducting a community-based, multi-country, pragmatic trial with the overall aim to compare the yield of detected TB cases and cost-effectiveness between two screening approaches, CAD4TB alone versus CAD4TB in combination with CRP.

Additionally, we will analyse household socioeconomic conditions and investigate if they correlate with TB diagnosis and TB treatment follow-up. We assess the initiation cost of TB treatment, and whether these medical activities cause financial struggles to households and the use of coping strategies such as borrowing and dissaving (the need to sell assets to pay for care).

The pragmatic community-based trial serves as a platform for multi-disease sub-studies:

<u>HIV and AHD sub-study:</u> Implementation and evaluation of the advanced HIV disease (AHD) care package in the community. For adults, the WHO defines AHD as having a CD4 cell count of <200 cells/µl or WHO stage 3 or 4 HIV [18]. The WHO recommends, since 2017, the implementation of an AHD care package, which has proven to reduce mortality in PWHIV with AHD, however this intervention remains poorly implemented [18–20]. The AHD care package consists of CD4 testing, and depending on the results and the setting, further screening for opportunistic infections such as TB and cryptococcal meningitis, and consequent (prophylactic and pre-emptive) treatment, including rapid ART initiation [18]. In our trial, we evaluate the use of the AHD care package during the community-based health campaign using a mixed-method approach, which includes studying acceptability and feasibility. We will also evaluate the diagnostic accuracy of VISITECT^®^ CD4 Advanced Disease (Accubio, UK; VISITECT CD4) in comparison with PIMA CD4 analyser (Abbott, USA; PIMA).

<u>Noncommunicable diseases (NCD) sub-study:</u> Screening for hypertension (HTN) and diabetes mellitus (DM). HTN is the most important cardiovascular risk factor and is increasing in sub-Saharan Africa [21]. There is similarly a dramatic increase in type 2 DM in these countries, attributed to urbanisation, epidemiological transitions and changes in lifestyle [21,22]. This screening is an important health service for an underserved population and provides important information on prevalence, risk factors and concurrent morbidities in the region.

## Methods and analysis

### Objectives

The primary objective of the trial is to investigate the hypothesis that a community-based active case finding strategy using CAD4TB with CRP (*CAD4TBv7-CRP approach)* screening will be non-inferior compared to CAD4TB screening alone (*CAD4TBv7 approach)* with regard to effectiveness, i.e. yield of detected TB cases, and cost-savings. Secondary objectives include i) assessing the cost-effectiveness of the two TB screening approaches against passive case finding, ii) assessing the households’ health expenditures to access TB treatment, ii) comparing household socioeconomic conditions between actively detected TB cases and passively detected TB cases, and to households with no TB diagnoses, and iv) modelling the counterfactual yield had active case finding not taken place to project the potential impact of broad implementation of the two approaches compared to passive case finding. Additional objectives include i) screening for HIV and AHD, hypertension and diabetes mellitus, ii) assessing the feasibility of implementing the AHD care package during a community-based health campaign, and iii) establishing a digital chest X-ray image collection for future validation of CAD software solutions for the detection of TB.

### Study design

This is a prospective, community-based, diagnostic, pragmatic trial in Lesotho and South Africa.

For efficiency reasons, the trial uses a paired screen-positive design to study the relative accuracy of two screening approaches tested in the same individual [23]. All participants receive a posterior-anterior digital chest X-ray which is analysed by CAD4TB version 7 (CAD4TBv7; Delft Imaging Systems, NL). In South Africa, where communities are easily accessible, a stationary X-ray system is used (FUJIFILM FDR Smart (FujiFilm, Japan) with a FXRD 1717V panel detector), while the terrain is more difficult in Lesotho and communities are more remote, requiring a more flexible, portable system (battery-operated portable Delft Light machine (Delft Imaging System, NL) with Canon CXDI-701 wireless flat panel detector).

The two screening approaches are shown in Figure 1. For the evaluation of the *CAD4TBv7 approach*, participants with a CAD4TBv7 score below or equal a pre-defined threshold (t1) are considered TB screen-negative, while participants with a CAD4TB score above t1 undergo confirmatory testing using Xpert MTB/RIF Ultra (Xpert Ultra; Cepheid, USA). For the *CAD4TBv7-CRP approach*, participants with a CAD4TBv7 score below or equal a pre-defined threshold (t2) are considered TB screen-negative. If the CAD4TBv7 score is within a defined threshold window (i.e. > t2 and ≤t3), the participants undergo additional point-of-care CRP testing (LumiraDX Limited, UK) from a capillary blood sample. In the *CAD4TBv7-CRP approach*, a confirmatory Xpert Ultra test will be conducted if the CRP result is above a pre-defined threshold or if CAD4TBvs7 is above the t3 threshold. If CRP result, is equal or smaller than its defined threshold the participant is also considered TB screen-negative.

**Figure 1.**
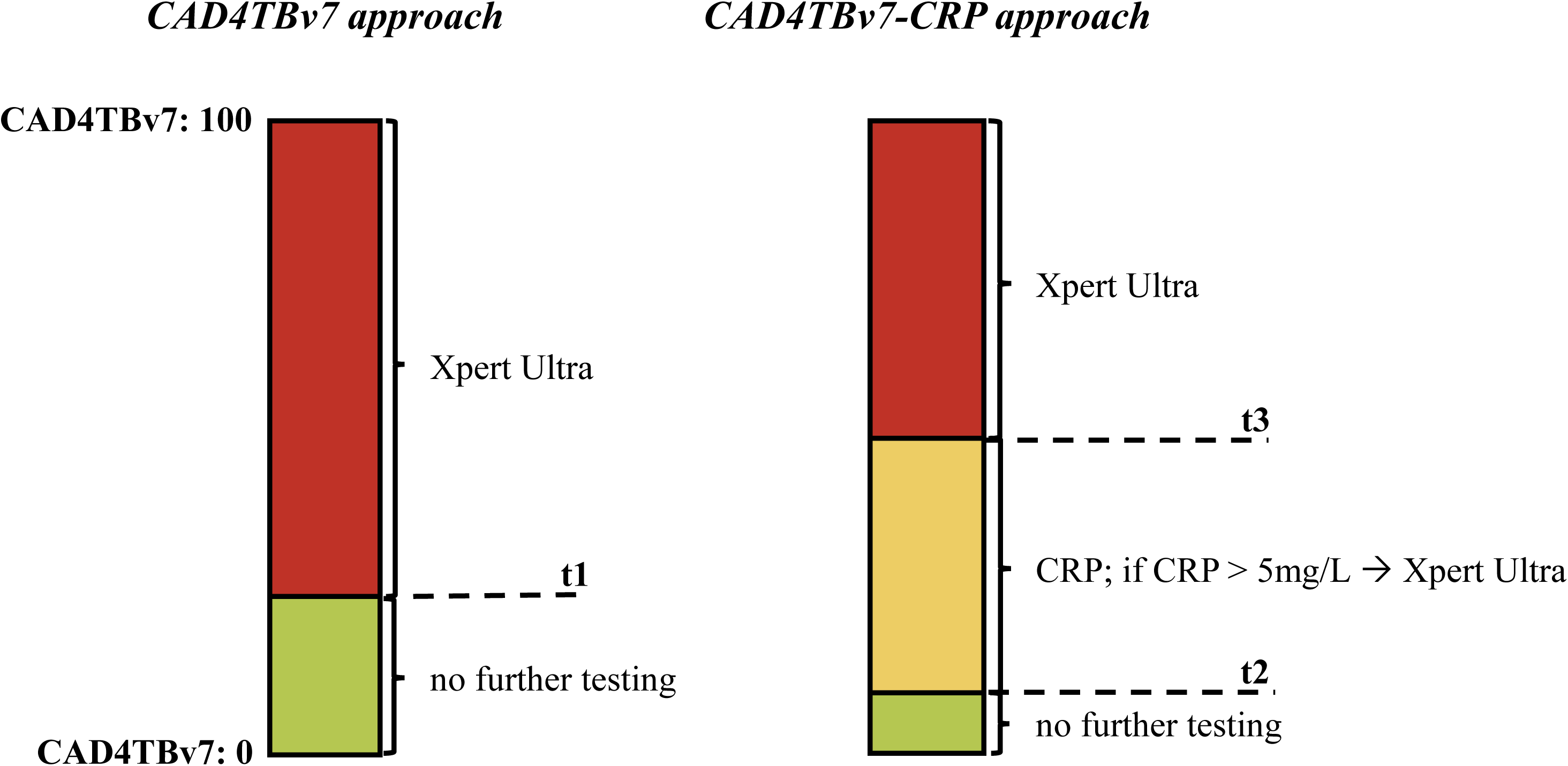
CAD4TBv7 thresholds for *CAD4TBv7* and *CAD4TBv7-CRP approaches* for TB screening.

The screen-positive design for testing two screening approaches with independent screening algorithms in the same individual leads to a partially overlapping consequential logic of the two approaches (e.g. an elevated CAD4TBv7 score may either indicate confirmatory testing per *CAD4TBv7 approach*, or CRP testing per *CAD4TBv7-CRP approach),* which will be taken into account in the analysis.

### Threshold determination

Initially, CAD4TB thresholds were determined based on secondary analysis of data from community TB screening in Lesotho, namely the TB Lesotho Prevalence Survey 2019 [24,25] because of similarity in population characteristics. Briefly, digital chest X-ray images from this external dataset were processed with CAD4TBv7 and the sensitivity and specificity of CAD4TBv7 were studied at integer thresholds against a reference of culture or Xpert Ultra positive (trace excluded) as previously described [25].

The selection of the thresholds was based on the following criteria:

**t1 threshold:** A CAD4TBv7 cut-off with a sensitivity of at least or close to 90% to meet the WHO for new TB diagnostic tests. For a screening test, the 90% sensitivity criterion is considered to be more important than 70% specificity.

**t2 threshold:** A CAD4TBv7 cut-off with sensitivity at least or close to 95% was chosen. If a 90% sensitivity can be achieved with CRP and the majority of true TB cases is above the upper CAD4TBv7 threshold, this would result in approximately 90% sensitivity overall for the CAD4TBv7-CRP screening.

**t3 threshold:** A CAD4TBv7 cut-off with a specificity at least or close to 95% was chosen. The upper threshold is designed to limit the number of required Xpert Ultra tests while still having an acceptable sensitivity.

After enrolment of 3,650 participants, for whom CAD thresholds derived from the Lesotho prevalence survey data were applied, it became apparent that individual site thresholds were required as the distribution of CAD scores is, as will be described elsewhere, highly dependent on the radiological equipment used. To account for these differences, we used datasets from community-based screening surveys or studies using similar radiological equipment and with similar CAD score distributions to each of the study sites to determine revised thresholds. Our data collected in South Africa showed a CAD4TBv7 score distribution very similar to the Lesotho TB prevalence survey data [24,25]. CAD4TBv7 score distributions of the Lesotho dataset were more similar to a community-based multi-morbidity survey conducted in KwaZulu-Natal, South Africa, the Vukuzazi study [26]. Therefore, the Vukuzazi data was used to determine the thresholds for Lesotho, while the Lesotho TB prevalence survey was used as a reference to determine the South Africa thresholds. The resulting CAD4TBv7 thresholds used in this study and their respective specificity and sensitivity for the used datasets are shown in Table 1.

**Table 1:** Diagnostic accuracy at the selected CAD4TBv7 thresholds for Lesotho and South Africa. Diagnostic accuracy was calculated for Lesotho using the Vukuzazi [26] dataset and for South Africa using the Lesotho TB prevalence survey data [24,25]. Accuracies are indicated for selected thresholds in the respective datasets (95% confidence intervals in brackets).

| **Lesotho** | **CAD4TBv7** | **Sensitivity** | **Specificity** |
| --- | --- | --- | --- |
| t1 | 25 | 88.1% [77.3% - 94.3%] | 72.6% [71.7% - 73.5%] |
| t2 | 10 | 95.5% [86.6% - 98.8%] | 47.2% [46.2% - 48.2%] |
| t3 | 50 | 61.2% [48.5% - 72.6%] | 95.0% [94.5% - 95.4%] |
| **South Africa** | **CAD4TBv7** | **Sensitivity** | **Specificity** |
| t1 | 13 | 90.1% [84.0% - 94.1%] | 74.2% [73.6% - 74.9%] |
| t2 | 5 | 93.7% [88.2% - 96.7%] | 54.2% [53.4% - 54.9%] |
| t3 | 44 | 70.4% [62.3% - 77.4%] | 94.9% [94.6% - 95.3%] |

For the 2,280 participants enrolled in Lesotho who were examined using CAD4TB thresholds before the revised thresholds were implemented, the new thresholds will be applied retrospectively reconstruct the screening algorithms.

The **CRP threshold** of 5 mg/L was determined based on existing scientific literature, the WHO recommendation on TB screening for adults and adolescents living with HIV [27], CRP data from a diagnostic accuracy study involving both CAD4TBv7 and CRP [28], and expert advice. A conservative selection of the CRP threshold will allow us to calculate the hypothetical yield of detected TB cases should another, higher threshold (like 10 mg/L) be used in the screening algorithm.

### Outcomes

#### Co-Primary outcomes – Effectiveness and cost-effectiveness

(1) Yield of detected TB cases per screening approach: number of positive Xpert Ultra results per screening approach

and

(2) Cost-effectiveness of the *CAD4TBv7-CRP approach* using the *CAD4TBv7 approach* as a comparator: the costs for each positive Xpert Ultra case detected in the *CAD4TBv7-CRP approach* compared to the *CAD4TBv7 approach*.

#### Secondary outcomes

- Extension of primary outcome: Cost-effectiveness of the *CAD4TBv7-CRP approach* and the *CAD4TBv7 approach* compared to passive case finding: cost per TB case detected.
- Equity analysis: 1. Socioeconomic status of households with actively detected TB cases compared to those with passively detected TB cases and to households with no TB cases: wealth quintiles distribution, income distribution and main source of income; 2. Average out of pocket payments (OOP) associated with accessing TB treatment for households with identified TB cases across different wealth quintiles.
- Health impact: The expected number of identified TB cases via passive case finding if the active case finding campaign had not taken place will be compared to the number of cases identified in the campaign.

Details regarding the secondary outcome measures and respective methodology will be presented in the respective trial reporting publications.

### Setting

The study is conducted in Butha Buthe District in Lesotho and in the Greater Edendale area of Msunduzi Municipality (uMgungundlovu District) in KwaZulu-Natal, South Africa (Figure 2). In 2022, Lesotho had an estimated TB incidence of 661 per 100,000 population [3]. During the same year, the HIV prevalence among persons aged between 15 and 49 years was 19.3% [29]. Butha Buthe District is characterized by mostly rural settings with an estimated population of 123,000 in 2021 [30], mainly subsistence farmers and mine workers as well as construction or domestic labourers who work in neighbouring South Africa. In South Africa, the estimated incidence of TB in 2022 was 468 per 100,000 population [26], and the HIV prevalence among 15 to 49–year-olds was 17.8% [29]. The Msunduzi Municipality with a population of around 218’000 people in 2022 [31] has high rates of both infectious diseases such as HIV (24.3% prevalence (people aged 15-49 years) in uMgungundlovu District [32]) and TB, as well as non-communicable diseases.

**Figure 2.**
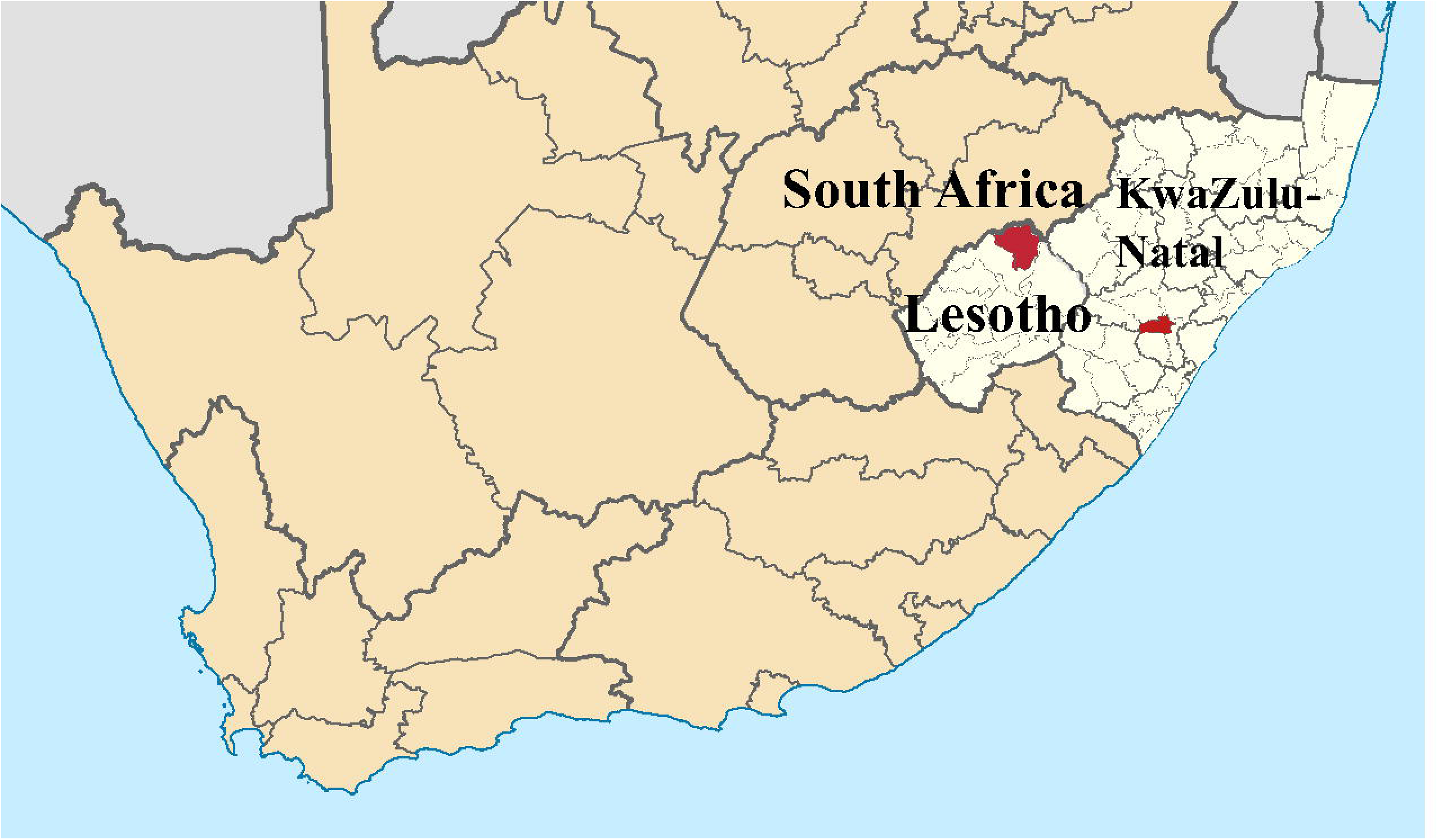
Butha Buthe District, Lesotho and Msunduzi Municipality n KwaZulu-Natal, South Africa, where the study is conducted (highlighted in red). Adapted from Wikimedia Commons (original maps by NordNordWest and Htonl) CC BY-SA 4.0. https://commons.wikimedia.org/wiki/File:Butha-Buthe_in_Lesotho.svg, https://commons.wikimedia.org/wiki/File:Map_of_South_Africa_with_uMgungundlovu_highlighted_(2011).svg, https://commons.wikimedia.org/wiki/File:Map_of_KwaZulu-Natal_with_Msunduzi_highlighted_%282011%29.svg.

A list of villages/neighbourhoods that are considered as sampling units within the Butha Buthe District and Greater Edendale area of Msunduzi Municipality in KwaZulu-Natal study area was compiled by the site teams together with local authorities and community leadership based on the following criteria:

1. the village/neighbourhood has at least 30 households (according to national census data)
2. no recent (past year), planned or on-going community-based health campaigns in this area during the trial
3. region with high TB incidence (>300/100,000 per year)
4. established collaboration with local authorities and the national TB programs

Villages/neighbourhoods were selected at random from this list by an independent statistician using a computer program (Stata). The sequences for visiting the selected villages/neighbourhoods is based on readiness for community entry and convenience. For each sampling unit, the number of households in the village/neighbourhood, the district of the village/neighbourhood, and the accessibility (average travel time and distance) to the corresponding health facility are recorded. Global Positioning System (GPS) coordinates are captured from each household (QGIS 3.28.11-Firenze). Each household within the selected village/neighbourhood is enumerated.

### Study participants

#### Community mobilization

The purpose of the TB screening campaign is explained to the village authorities/community advisory board to obtain their approval. Once these authorities approve the campaign, they inform the community about the date and purpose of the campaign. Approval for the campaign is also sought from the local health authorities. Community mobilisation activities may include community dialogues, loud hailing and motorcades that stop at community hotspots to talk with and inform people about the study. Pamphlets and posters explaining the study and social media may also be used as part of the study’s community mobilisation strategies. On the day of a campaign, a team consisting of lay-counsellors, nurses and drivers arrives early at the village chief’s place of residence to formally ask again for permission to start the campaign. The study team then parcels out the village and organizes the team ensuring that each household will be visited and equipment has been set up. Community mobilisers recruited from each of the study areas then move door to door in their community introducing the study to the head-of-household, or their representative, after asking for permission to enter the household.

#### Eligibility

Participants eligible for participation in this trial are people aged 18 years or older. Participants who are seriously ill and participants with any condition for which participation in the study, as judged by the investigator or a designated staff member, could compromise the well-being of the participant or prevent, limit or confound protocol specified assessments are excluded from the study. Furthermore, participants currently on anti-TB treatment are excluded (preventive TB treatment permissible).

In Lesotho, self-reported pregnancy constitutes an additional exclusion criterion.

Persons found to need immediate medical care are referred to the nearest health facility.

#### Participant information and consent

Informed consent is obtained from all persons enrolled in the study. Verified translations of consent forms have been developed in the local languages, Sesotho and Zulu. The information session is held at the participant’s home (Lesotho) or in a private environment around the study site within the community (South Africa). Written consent is obtained with the signature of the informed consent form. If the participant is illiterate, an impartial witness must be present during the entire information session. The facilitator explains to the participant the information contained in the written document and asks whether they give their consent to participate. The participant’s voluntary consent is documented with their fingerprint on the consent form. Participants may decide not to continue participating in the study at any time for any reason if they wish to do so without any particular involvement, but they are informed that the data collected up to the time of withdrawal will be included in the analysis. The investigator may decide to terminate the study prematurely for reasons of ethical concerns, insufficient participant recruitment or changes in accepted clinical practice that make it inappropriate to continue the study.

### Study procedures

Table 2 presents a summary of all measurements conducted in the study.

**Table 2:**
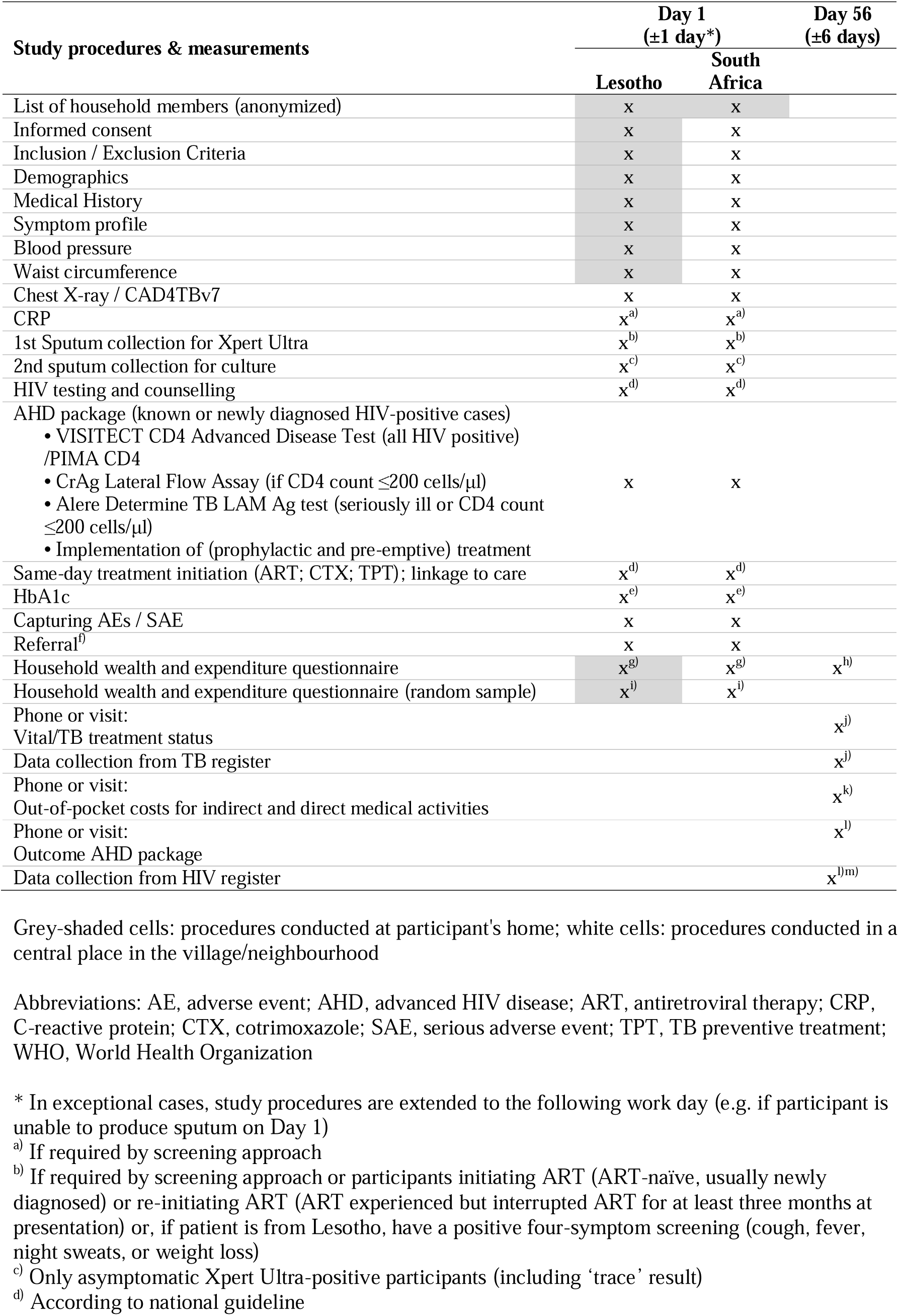
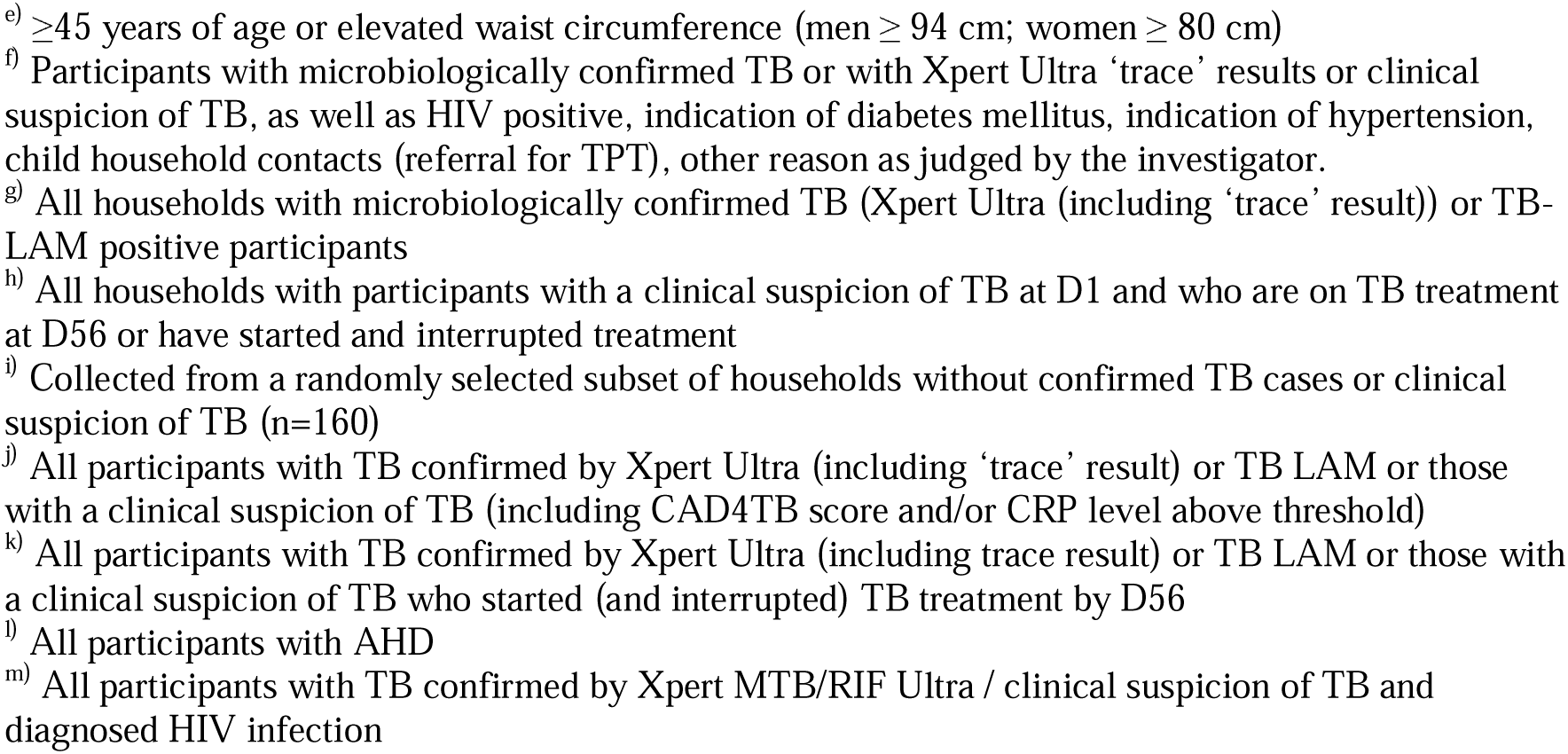
Overview of study procedures and measurement.

Depending on the size of the sampling unit, villages/neighbourhoods are visited over a period of one or several days after involving the communities and their representatives as described under community mobilization. For participating households, aggregate indirectly-identifiable, non-sensitive data (age and gender, relationship to household head, locator information) will be collected from the household head. Interested household members are introduced to the study team members for recruitment, the process of informed consent and assessment of eligibility. Study procedures start either in the households (e.g. informed consent, interviews) and/or at a central or convenient place in the community (e.g. X-ray, point-of-care testing; Table 2). However, the exact setup, which defines sequence and location of the procedures, is adapted to different local circumstances, always respecting confidentiality and privacy. For all eligible participants, we collect demographic information and establish the TB symptom profile and the participant’s medical history.

#### TB screening

A posterior-anterior digital chest X-ray is acquired for all participants and analysed by CAD4TBv7. A trained and qualified person in compliance with national regulations operates the digital chest X-ray machine. Participants are assigned to study procedures according to both the *CAD4TBv7* and the *CAD4TBv7-CRP approach* following the paired screen-positive design as described in detail under Study design and in Figures 1 and 3.

**Figure 3.**
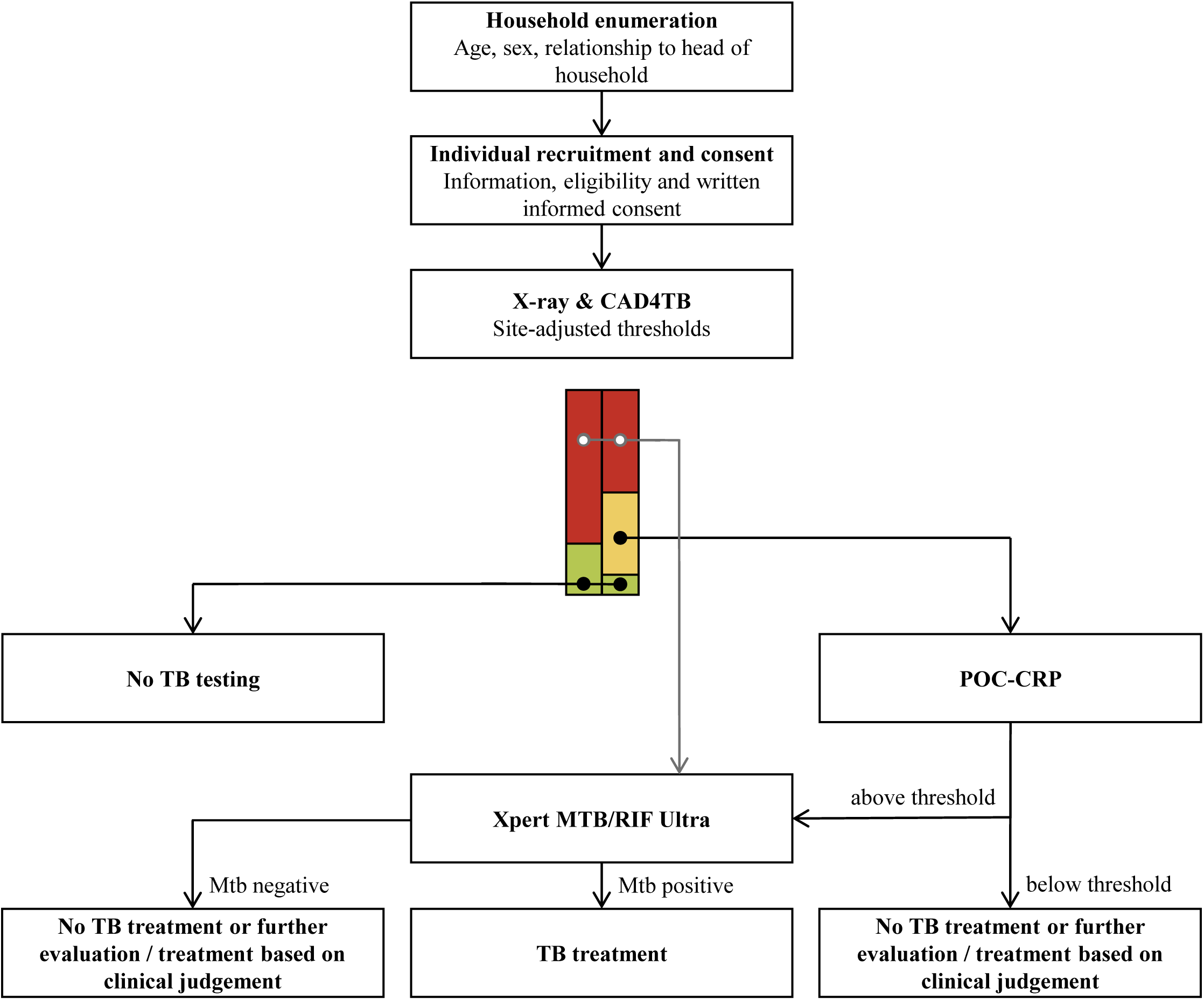
Test algorithm for *CAD4TBv7* and *CAD4TBv7-CRP* TB screening approaches. Figure 3 shows the screening approaches presented in more detail in Figure 1.

Participants eligible for Xpert Ultra testing are given instructions to explain how to produce a quality spot sputum sample. Xpert Ultra testing is done within 24 hours. In case of a positive test result, participants receive a referral letter documenting the test results and are referred to the nearest healthcare facility for treatment according to national guidelines in Lesotho and South Africa, respectively. Asymptomatic participants with a positive Xpert Ultra result are asked to produce a second sputum sample for liquid mycobacterial culture (MGIT; BACTEC MGIT 960 TB System, Becton Dickinson, USA) at a central laboratory. The result is made available to the referral healthcare facility. Screening test negative participants with a clinical suspicion of TB (TB signs/symptoms or radiological findings) are referred to the nearest health facility for further investigations, while screening test negative participants with a low clinical suspicion of TB are asked to visit the health facility if they have any TB symptom which persists for more than 4 weeks. Participants who cannot produce sputum on day 1 and day 2 are referred to the health facility for further investigations if Xpert Ultra is indicated by either of the screening approaches. A ‘trace’ result by Xpert Ultra is not considered a positive confirmatory test for TB. Participants with an Ultra ‘trace’ result are referred to the health facility to be further investigated and managed according to national guidelines in Lesotho and in South Africa.

Fifty-six (±6) days after screening, the study team calls or visits the following participants to assess vital and TB treatment status: i) those with TB confirmed by Xpert Ultra, ii) those with an Ultra ‘trace’ result, iii) those who are TB LAM positive (see HIV and AHD sub-study), iv) those who are MGIT positive, v) those with a clinical suspicion of TB, and vi) those for whom an Xpert Ultra was indicated based on CAD4TBv7 and/or CRP results but who were unable to produce sputum and were therefore referred. In addition, data are collected from the HIV register for participants living with HIV if the participant has TB confirmed by Xpert Ultra or clinical suspicion of TB.

For the same participants, the study team collects details about any confirmatory TB diagnosis and TB treatment status from National TB Programme (NTP) registers or TB Clinic Register.

#### Households’ socioeconomic and expenditure data

We collect information on the household wealth and health expenditures of all households in which at least one member tested positive for TB by Xpert Ultra (including trace result) and/or TB LAM (see HIV and AHD sub-study); households with at least one member with clinical suspicion of TB at D1 or who could not produce sputum, but is on TB treatment at D56; and a randomly selected subset of households without any participants testing positive for tuberculosis (n=160). Household members are interviewed using structured questionnaires. To compare the socioeconomic profiles of TB cases from active and passive case finding, we include adults (n=150) who present to health facilities within the study area and are newly diagnosed with TB within the past month through passive case finding.

Information on medical and non-medical out-of-pocket payments and indirect costs of seeking care (in terms of work days lost) as well as any coping strategies used (e.g. borrowing, selling assets) to pay for care are collected at Day 56 from participants with TB confirmed by Xpert Ultra (including trace) or TB LAM and those with a clinical suspicion of TB or who could not produce sputum and started TB treatment.

#### HIV and AHD

Point of care (POC) tests are available to facilitate the implementation of the AHD care package, including the semi-quantitative VISITECT CD4, the urine Alere Determine TB-lipoarabinomannan (Abbott, USA), and CrAg lateral flow assay (Immy, USA). The AHD care package is offered to eligible persons as indicated in Table 3.

**Table 3:** Overview of advanced HIV care package* offered to eligible individuals aged 18+ years during the community-based TB screening campaign.

|  | **Intervention** | **Eligibility** |
| --- | --- | --- |
| **Screening & diagnosis** | HIV testing & counselling<br>CD4 cell count<sup>1)</sup><br>LF- LAM screening<sup>2)</sup> | Per national guidelines<br>HIV positive<br>CD4 ≤ 200 cells/μl or seriously ill |

| | CrAg screening<sup>3)</sup> | CD4 ≤ 200 cells/μl |
| --- | --- | --- |
| **Prophylaxis / pre-emptive treatment** | Cotrimoxazole prophylaxis<br>TB preventive treatment | Per national guidelines |
| **Same-day ART initiation** | ART initiation | Per national guidelines |

Abbreviations: ART, antiretroviral therapy; CrAg, cryptococcal antigen; LF-LAM, lateral flow lipoarabinomannan assay; TB, tuberculosis; WHO, World Health Organization.
\* Adapted from WHO [18]
<sup>1)</sup> Until April 2023: VISITECT CD4 (Accubio, UK); after April 2023: PIMA CD4 (Abbott, USA)
<sup>2)</sup> TB LAM (Abbott, USA)
<sup>3)</sup> CrAg (IMMY, USA)

After a VISITECT CD4 batch recall due to a suboptimal specificity in April 2023, PIMA CD4 replaced VISITECT CD4. In a subgroup (n= 609) of consequently recruited PWHIV in South Africa, VISITECT and PIMA were used in parallel to determine the diagnostic accuracy of VISITECT CD4. In order to enable same-day antiretroviral therapy (ART) initiation, participants may have to undergo TB testing by Xpert Ultra as per the national HIV guidelines. Such additional Xpert Ultra testing in AHD patients increases the diagnostic yield for TB screening and such test results will not be included in the primary analysis.

Participants who are eligible to start same-day ART, cotrimoxazole prophylaxis, and/or tuberculosis prevention therapy according to national guidelines receive medication for 2 to 4 weeks and are referred to the nearest health facility to link to care and be registered as per national guidelines. If the patient wishes, treatment can also be started at the facility.

Pregnant women with HIV infection are referred to an antenatal care clinic.

All participants who were diagnosed having AHD are followed up by a call or visit after 56 days to collect information on vital status and on treatment and outcome indicators. Similarly, information is collected from the HIV registers.

#### NCD screening

The screening algorithm for DM and HTN is shown in Table 4.

**Table 4:**
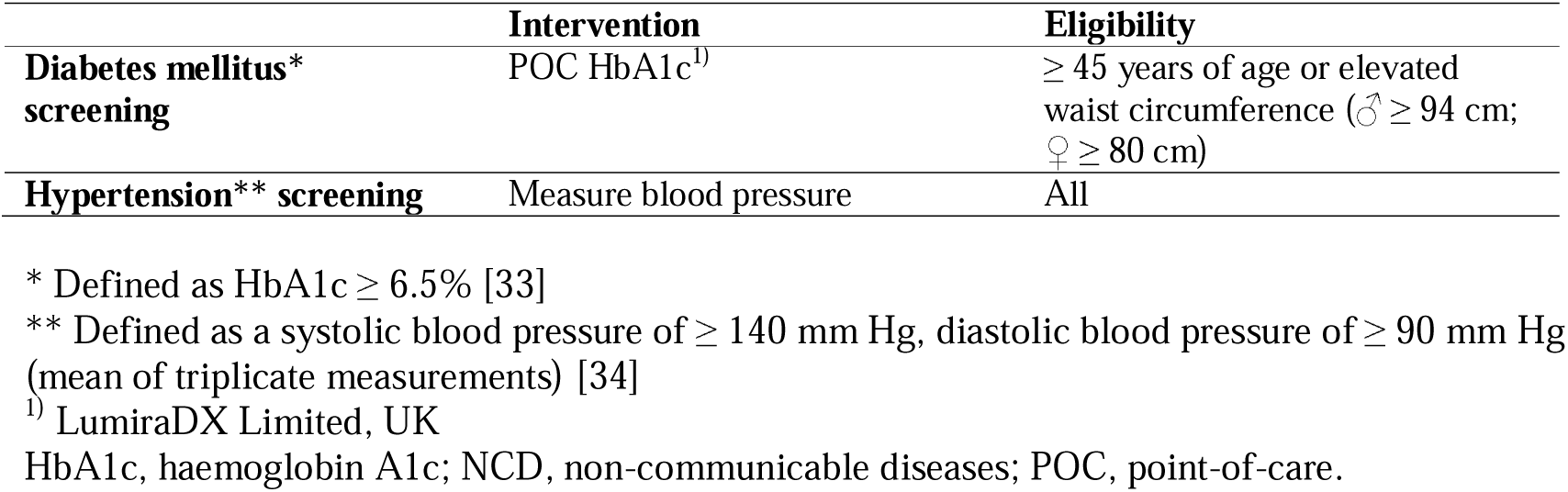
Overview of NCD screening offered during community-based TB screening campaign. Diabetes mellitus and hypertension screening is offered to all enrolled participants fulfil the below eligibility criteria.

Participants with presumptive DM or HTN receive a referral letter documenting the findings and are referred to the nearest healthcare facility for further evaluation.

#### Intervention costs data

For the costing of the active-case finding campaign, detailed cost data for implementing both screening approaches are systematically collected throughout the trial. Intervention costs are gathered from a program and healthcare perspective using cost collection tools administered to the trial staff [35]. Resource use is measured by reviewing the implementation of the intervention, direct observation, and interviews with trial implementation staff. The data collection is supplemented with information from document review and secondary sources. Both financial and economic costs are estimated, with economic capital costs being discounted at an annual rate of 3% [36]. However, different discounting rates will be used in the sensitivity analysis.

#### Study tools

Clinical study data as well as the household socioeconomic and expenditure data of screened study participants and TB patients identified through passive case finding is collected and managed using REDCap electronic data capture tools hosted at the Swiss TPH [37,38]. The tool contains a computer-generated time stamped audit trail. Data from TB patients identified through passive case finding are kept in a separate REDCap database.

Household enumeration data is collected on paper forms and transcribed into REDCap.

The costing tool captures the site-specific resources used for the intervention and their respective unit costs by cost category in Microsoft Excel.

#### Data management and study monitoring

Data is collected by a trained study team using either password-protected portable electronic devices (tablets) with incorporated REDCap data entry forms (electronic data capture) or on validated structured paper-based forms, as indicated above. Data is managed and stored in accordance with local and national information governance specifications. Project data is handled with utmost discretion and is only accessible to authorized personnel who require the data to fulfil their duties within the scope of the research project. To ensure confidentiality in all data collection tools, participants are only identified by a unique participant number, which will be anonymized and cannot be traced back to the person without a separate ID key which is safely stored on site. Data is regularly reviewed by central and local data managers. Queries regarding inconsistencies and missing data are raised within REDCap. The Swiss Tropical and Public Health Institute’s Clinical Operations Unit is responsible for trial monitoring in Lesotho and South Africa. A risk-based monitoring approach is chosen for this study, taking the study’s risk assessment [39] into consideration. On-site monitoring is scheduled at month 1 and month 3 of study initiation and in six-monthly intervals thereafter and consists of a systematic examination of study-related activities and documents.

### Sample Size

The sample size calculation is based on extensive simulations with accuracy estimates from various studies using CRP, CAD4TB and Xpert Ultra in high TB burden settings, taking into account HIV status, history of TB, symptoms, and realistic conditional dependence of the different tests [40–43]. We choose a non-inferiority margin of 10%. The sample size of 20,000 participants gives an 80% probability that the lower bound of the one-sided 95% confidence interval of the difference in the yield of TB cases between the two screening approaches will exceed this margin. The sample size was estimated using an asymptotic approach with the prop.test function of the R software (R 4.0.3. R Foundation for Statistical Computing, Vienna, Austria). As both screening approaches will be observed in each person, the calculation is a conservative estimate and the true power is likely higher than 80%.

### Statistical analyses

Statistical analyses will be performed using Stata [44] and R software. Baseline characteristics will be described using summary statistics such as median and interquartile range or number and percentage. Yield of TB cases will be described by screening approach using summary statistics. For the non-inferiority comparison of the TB yield, confidence intervals (CI) will be estimated using an appropriate model [45,46], accounting for the paired nature of the observations. Primary analyses will be performed on a complete case, per protocol set, with no imputation for missing data. Any missing data for one individual that will not allow for the assessment of the outcome for one of the approaches will lead to the exclusion of the individual from the analyses. If non-inferiority is demonstrated, then we will assess for superiority following intention to test principles. Effect modification of the primary outcome by site and HIV status will be assessed by stratification, acknowledging that the power will be low.

For the cost-effectiveness analysis, we will estimate the costs per additional case detected if the *CAD4TBv7-CRP approach* proves to be superior to the *CAD4TBv7 approach*. If the *CAD4TBv7-CRP approach* is not superior, the two approaches will be compared based on the average cost per case detected. In the former case, we will calculate an incremental cost-effectiveness ratio (ICER) and assess the uncertainty with cost-effectiveness acceptability curves, showing the probability that the intervention is cost-effective assuming different willingness to pay thresholds. In the latter scenario, we will conduct a probabilistic sensitivity analysis to explore the impact of parameter uncertainty on the cost per case.

For the household socioeconomic analyses, missing data will be assessed in two steps. Participants who have been lost to follow-up will be compared using univariate analysis to patients whose data are complete. Similarly, participants who decline participation in the household wealth and expenditure surveys will be compared based on basic demographics and disease status with those who participate. The first step is in order to understand if data are missing at random. Should the patients not be missing at random, then multiple imputation methods will be explored to estimate the values of these patients [47].

### Harms

Overall, the community-based comparison of the two screening approaches using CAD4TBv7 and CRP is considered low risk to trial participants. Likewise, the use of routine diagnostic tests for HTN and DM as well as of tests within the AHD care package within the community constitute a minimal risk. The potential benefits of screening participants for these diseases likely outweigh any small risks. As for any clinical study, there is a possibility of unknown and unforeseen risk; that possibility is small for this study.

Safety information is collected by the study team on the day(s) of the community-based study procedures. This information will be obtained by solicitation from, observation of and/or reporting by the study participant. Safety information includes adverse events and serious adverse events as well as incidental findings.

### Patient and public involvement

This study uses existing community engagement structures to work collaboratively with community groups following good participatory practice guidelines. This is accomplished in community advisory board meetings, in smaller assemblies in the recruitment areas and through direct communication with local stakeholders, including the TB Program Manager, Regional and District Medical Officers or representatives of the City Council.

### Trial oversight

An independent scientific advisory board (SAB) consisting of experts in TB/HIV clinical research, respiratory medicine, innovative diagnosis and care models, medical statistics and epidemiology was set up for this trial. The SAB meets yearly to review the progress of the trial and provides scientific and strategic advice.

### Study schedule and trial status

The first participant was enrolled in September 2022. Enrolment will continue until September 2024. At the time of manuscript submission, 20,006 study participants have been screened in the two countries. The final D56 follow up visit is anticipated to be completed in November 2024.

### Ethics and dissemination

Ethics approval for the trial was obtained from the National Health Research Ethics Committee in Lesotho (NH-REC, ID52-2022), the Human Sciences Research Council Research Ethics Committee (HSRC REC, REC 2/23/09/20) and the Provincial Health Research Committee of the Department of Health of KwaZulu-Natal (KZ_202209_022) in South Africa and from the Swiss Ethics Committee Northwest and Central Switzerland (EKNZ, AO_2022-00044).

The study is conducted in compliance with the current version of the Declaration of Helsinki, ICH-GCP E6(R2) as well as all the national legal and regulatory requirements

The results of the study will be shared through peer-reviewed articles, conference presentations as well as through communication offices of the consortium partners and the project’s website (https://tbtriage.com/). Dissemination to local and national health authorities will be ensured, for example through dissemination meetings and symposia.

## Discussion

This manuscript describes a diagnostic pragmatic trial designed to test and compare the relative effectiveness and cost-effectiveness of community-based TB screening algorithms. To our knowledge, this is the first comparative evaluation of two screening approaches for pulmonary TB using CAD4TBv7 alone or in combination with point-of-care CRP. Results from this study will help in identifying the most (cost-)effective strategy and will inform national policy on TB screening approaches.

A previous trial within the TB TRIAGE+ project assessed the diagnostic accuracy of CAD4TBv7 and CRP in adults presenting with one or more cardinal tuberculosis symptoms (cough, fever, night sweats, loss of weight) at health facilities in Butha Buthe and the Greater Edendale area in KwaZulu-Natal. While CAD4TBv7 was close to meeting the WHO TPP criteria for a TB screening test of 90% sensitivity and 70% specificity in this population, CRP had a similar sensitivity but a much lower specificity [28]. While CRP may not be suitable as a stand-alone screening test, its use in a sequential positive serial screening algorithm in a community screening program together with CAD4TBv7 like in our *CAD4TBv7-CRP approach* may still be warranted as it could increase the pre-test probability before confirmatory testing.

A recent review and analysis of 23 national and 5 subnational TB prevalence surveys showed that 50.4% of confirmed TB was subclinical, defined as negative at symptom screening, but confirmed on bacteriological testing [48]. Similar proportions of subclinical TB were reported from the TB prevalence survey from Lesotho and South Africa [24,26] and a screening campaign in KwaZulu-Natal [49]. The high prevalence of subclinical TB requires a screening approach, which is not based on initial symptom screening, because symptom-based screening would miss half of all infectious TB cases. Our study therefore evaluates symptom-agnostic screening algorithms.

The trial applies a paired screen-positive design. This study design allows for higher statistical power and minimizes the variability compared to other randomized trial designs. In the analysis, the *CAD4TBv7 approach* and *CAD4TBv7-CRP approach* screening algorithms will be compared using outcomes that would have occurred under each approach within the same participant. From a participant perspective, this design may possibly result in needing to undergo a higher number of tests compared to other designs.

Embedded in this trial is a multi-disease sub-study. Provision of an AHD care package has proven to reduce mortality in PWHIV with AHD and is recommended by the WHO. However, this intervention remains poorly implemented [18–20]. We will therefore evaluate the feasibility to implement an AHD care package in the community during the trial. NCDs, such as diabetes and hypertension cause a large burden of morbidity and mortality in sub-Saharan Africa. For example, diabetes is a known risk factor for the initial development of TB and for the precipitation of active TB from latent infection. Since we are targeting remote communities with limited access to health care, it would be unethical to screen only for tuberculosis, which is a relatively rare disease even in these high disease burden areas. Therefore, we aim to provide screening for NCDs in the community and linkage to care at the nearest health facility. This activity also serves as a cross-sectional study of disease prevalence in regions where data on NCD is still very limited.

In summary, our study will improve access to quality health care for hard-to-reach populations in Southern Africa and (early) linkage to care. The development of an accurate, robust and cost-effective community-based TB screening algorithm as part of health campaigns will ultimately contribute to ending the epidemic of TB, as targeted by Sustainable Development Goal 3.

## Data Availability

This is a protocol manuscript without data

## Declarations

### Ethics approval and consent to participate

See Methods and Design section.

### Consent for publication

Not applicable.

### Availability of data and materials

Not applicable.

### Competing interests

The authors declare no competing interests.

### Funding

This work was supported by the European and Developing Countries Clinical Trials Partnership (EDCTP) as part of the TB TRIAGE+ project (grant number RIA2018D-2498). The funders had no role in study design, data collection and analysis, decision to publish, or preparation of the manuscript.

### Authors’ contributions

All co-authors contributed to conceiving and designing the study. All authors reviewed the draft manuscript, and all contributed to and approved the final manuscript.

## Acknowledgements

The study team would like to sincerely thank all the participants in this study. We are furthermore very grateful to local and national health authorities who provided their support as well as to our study teams. We would like to thank Prof. Emily B Wong for sharing the Vukuzazi data as well as Dr. Llang B. Maama for the Lesotho TB prevalence survey data. Finally, we thank Profs. Anja van ‘t Hoog (in memoriam), Graeme Meintjes (University of Cape Town), Sian Floyd (London School of Hygiene and Tropical Medicine) and Christina Yoon (University of California, San Francisco) for important advisory input in the development of this research protocol. Finally, we sincerely thank Dr. Debora Bade from EDCTP for her support of the TB TRIAGE+ project.

## Notes

### Competing Interest Statement

The authors have declared no competing interest.

### Clinical Trial

NCT05526885

### Author Declarations

Ethics committees/IRBs of National Health Research Ethics Committee in Lesotho, Human Sciences Research Council Research Ethics Committee and the Provincial Health Research Committee of the Department of Health of KwaZulu-Natal in South Africa and the Swiss Ethics Committee Northwest and Central Switzerland gave ethical approval for this work.

